# Longitudinal immunological analyses reveal inflammatory misfiring in severe COVID-19 patients

**DOI:** 10.1101/2020.06.23.20138289

**Authors:** Carolina Lucas, Patrick Wong, Jon Klein, Tiago B.R. Castro, Julio Silva, Maria Sundaram, Mallory K. Ellingson, Tianyang Mao, Ji Eun Oh, Benjamin Israelow, Maria Tokuyama, Peiwen Lu, Arvind Venkataraman, Annsea Park, Subhasis Mohanty, Haowei Wang, Anne L. Wyllie, Chantal B.F. Vogels, Rebecca Earnest, Sarah Lapidus, Isabel M. Ott, Adam J. Moore, M. Catherine Muenker, John B. Fournier, Melissa Campbell, Camila D. Odio, Arnau Casanovas-Massana, Yale IMPACT Team, Roy Herbst, Albert C. Shaw, Ruslan Medzhitov, Wade L. Schulz, Nathan D. Grubaugh, Charles Dela Cruz, Shelli Farhadian, Albert I. Ko, Saad B. Omer, Akiko Iwasaki

## Abstract

Recent studies have provided insights into the pathogenesis of coronavirus disease 2019 (COVID-19)^1–4^. Yet, longitudinal immunological correlates of disease outcome remain unclear. Here, we serially analysed immune responses in 113 COVID-19 patients with moderate (non-ICU) and severe (ICU) disease. Immune profiling revealed an overall increase in innate cell lineages with a concomitant reduction in T cell number. We identify an association between early, elevated cytokines and worse disease outcomes. Following an early increase in cytokines, COVID-19 patients with moderate disease displayed a progressive reduction in type-1 (antiviral) and type-3 (antifungal) responses. In contrast, patients with severe disease maintained these elevated responses throughout the course of disease. Moreover, severe disease was accompanied by an increase in multiple type 2 (anti-helminths) effectors including, IL-5, IL-13, IgE and eosinophils. Unsupervised clustering analysis of plasma and peripheral blood leukocyte data identified 4 immune signatures, representing (A) growth factors, (B) type-2/3 cytokines, (C) mixed type-1/2/3 cytokines, and (D) chemokines that correlated with three distinct disease trajectories of patients. The immune profile of patients who recovered with moderate disease was enriched in tissue reparative growth factor signature (A), while the profile for those with worsened disease trajectory had elevated levels of all four signatures. Thus, we identified development of a maladapted immune response profile associated with severe COVID-19 outcome and early immune signatures that correlate with divergent disease trajectories.

## Introduction

Coronavirus Disease 2019 (COVID-19) is caused by severe acute respiratory syndrome coronavirus 2 (SARS-CoV-2), a highly infectious, zoonotic virus that exploits angiotensin-converting enzyme 2 (ACE2)^5,6^ as a cell entry receptor. SARS-CoV-2 has generated an ongoing global pandemic that reaches almost every country, linked to hundreds of thousands of deaths in less than 6 months. Clinical presentation of COVID-19 involves a broad range of symptoms comprising fever, fatigue, diarrhoea, conjunctivitis and myalgia, in addition to the respiratory-specific symptoms including dry cough, shortness of breath and viral pneumonia^3,7,8^. In severe cases, patients develop critical disease characterized by pneumonia that may be exacerbated by pulmonary edema, respiratory failure, cardiac damage, pulmonary emboli, thromboses, systemic shock, and multi-organ failure^3,9^. Understanding the nature of the immune response that leads to recovery over severe disease is key to developing effective treatment against COVID-19.

Coronaviruses, including Severe Acute Respiratory Syndrome (SARS-CoV) and Middle Eastern Respiratory Syndrome (MERS), typically induce strong inflammatory responses and associated lymphopenia^10,11^. Initial studies characterizing immune cells in the peripheral blood of COVID-19 patients have reported increases in inflammatory monocytes and neutrophils and a sharp decrease in lymphocytes^1–4^. Moreover, in parallel with observations of infections with MERS and SARS, an inflammatory milieu containing IL-1β, IL-6, and TNF-α has been associated with worse disease outcomes^1,2,4,8,12^. Despite these analyses, immune response dynamics during the course of SARS-CoV-2 infection and its possible correlation with clinical trajectory remain unknown.

Immune responses against pathogens are divided roughly into three types^13–15^. Type-1 immunity, characterized by T-bet-dependent responses and IFN-γ, is generated against intracellular pathogens including viruses. In type-1 immunity, pathogen clearance is mediated through effector cells including ILC1, NK cells, cytotoxic T lymphocytes, and Th1 cells. Type-2 immunity, which relies on the GATA-3 transcription factor, mediates anti-helminths defense through effector molecules including IL-4, IL-5, IL-13, and IgE designed to expel these pathogens through the concerted action of epithelial cells, mast cells, eosinophils, and basophils. Type-3 immunity, orchestrated by the RORγt-induced cytokines IL-17, IL-22 secreted by ILC3 and Th17 cells, is mounted against fungi and extracellular bacteria to elicit neutrophil-dependent clearance. In this study, we focused on the longitudinal analysis of these three types of immune responses to COVID-19 patients and identified correlations between distinct immune phenotype and disease.

## Results

### Overview of immunological features in COVID-19 patients

To determine immune phenotypes underlying disease trajectories in COVID-19, we conducted a longitudinal analysis of COVID-19 patients. One hundred and thirteen COVID-19 patients admitted to Yale New Haven Hospital (YNHH) between the 18th of March through the 27th of May 2020, were recruited to the Yale IMPACT study (Implementing <u>M</u>edical and <u>P</u>ublic Health <u>A</u>ction Against Coronavirus <u>CT</u>). We assessed a) viral RNA load quantified by RT-qPCR using nasopharyngeal swabs, b) levels of plasma cytokines, chemokines, and c) leukocyte profiles by flow cytometry using freshly isolated peripheral blood mononuclear cells (PBMCs). We performed 253 collections and follow-up measurements on the patient cohort with a range of one to seven longitudinal time-points that occurred 3-51 days post symptom onset. In parallel, we enrolled 108 volunteer healthcare workers (HCW) into the IMPACT study, whose samples served as healthy controls (SARS-CoV-2 negative by RT-qPCR and serology).

Basic demographic information stratified by disease severity is displayed in Extended Table 1. Hospitalized patients were stratified into moderate and severe based on oxygen levels and intensive care unit (ICU) requirement (Fig.1a). Among our cohort, patients who developed moderate or severe disease did not significantly differ with respect to age or sex. Body mass index (BMI) was generally increased among patients with severe disease, and extremes in BMI correlated with an increased relative risk of mortality (RR BMI ≥ 35: 1.62 [95% CI .81-3.22]) (Extended Table 1, Extended data Fig. 1a,b). Exposure to select therapeutic regimens of interest was assessed in both moderate and severe disease severities (Extended Data Fig 1c). Initial presenting symptoms demonstrated a preponderance of headache (54.55%), fever (64.47%), cough (74.03%), and dyspnoea (67.09%) with no significant difference in symptom presentation between moderate or those that eventually developed severe disease. As reported elsewhere, blunting of taste (hypogeusia) and smell (anosmia) were occasionally reported in both moderate and severe disease but demonstrated no significant difference between groups^16^. Finally, mortality was significantly increased in patients who were admitted to the ICU over those who were not (27.27% vs 3.75%; RR: 7.27 [95% CI 2.1 – 25.19], p-value < .001) (Extended Table1).

**Figure 1.**
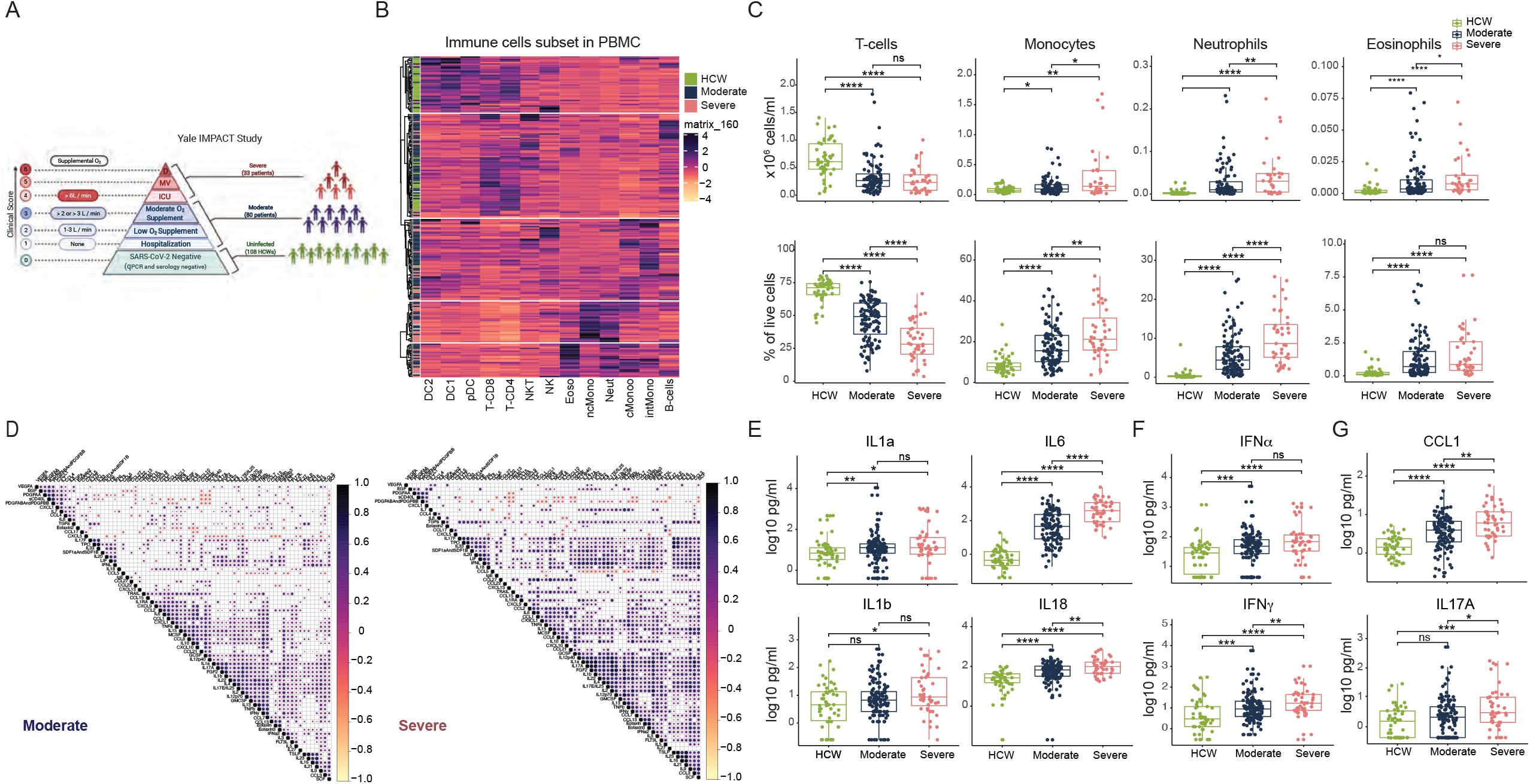
Overview of immunological features in COVID-19 patients. **a**, Overview of cohort, including healthy donors (health care workers-HCW), and COVID-19 moderate and severe patients enrolled. Ordinal scores assigned according to clinical severity scale. Non-infected donors (Clinical Score 0), moderate disease status (Clinical Score 1, 2 and 3): (1) SARS-CoV-2 infection requiring hospitalization, (2) infection requiring non-invasive supplemental oxygen (<3 L / min), (3) infection requiring non-invasive supplemental oxygen (>3 L / min). Severe disease status (Clinical score 4 and 5): (4) Intensive care unit (ICU) admission and supplemental oxygen (>6 L / min), (5) invasive mechanical ventilation (MV). Clinical Score 6: deceased. **b**, Heat map comparison of the major immune cell populations within peripheral blood mononuclear cells (PBMCs) in COVID-19 patients, graded as either moderate (n=121) or severe (n=43), or healthcare workers (n=43). Subjects are arranged across rows, with each coloured unit indicating the relative distribution of an immune cell population normalized against the same population across all subjects. K-means clustering was used to arrange patients and measurements. **c**, Immune cell subsets of interest, plotted as a concentration of millions of cells per mL of blood or as a percentage of live singlets. Each dot represents a separate time point per subject (HCW, n=50; Moderate, n=117; Severe, n=40). **d**, Correlation matrices across all time points of 71 cytokines from patient blood comparing moderate patients to severe patients. Only significant correlations are represented as dots. Pearson’s correlation coefficients from comparisons of cytokine measurements within the same patients are visualized by colour intensity. **e**, Quantification of prominent inflammatory cytokines, **(f)** interferons type I and II, and **(g)** CCL1 and IL-17 presented as Log10-transformed concentrations in pg/mL. Each dot represents a separate time point per subject (HCW, n=50; Moderate, n=117; Severe, n=40). Significance of comparisons were determined by Wilcoxon Rank-Sum Test and indicated as such: * p < 0.05, ** p< 0.01, *** p < 0.001, and **** p <0.0001.

We analysed PBMC and plasma samples from moderate and severe COVID-19 patients and healthy HCW donors (Fig. 1a) by flow cytometry and ELISA to quantify leukocytes and soluble mediators, respectively. An unsupervised heatmap was constructed from the main innate and adaptive circulating immune cell types; this analysis revealed marked changes in COVID-19 patients compared to uninfected HCW (Fig. 1b). As recently reported ^1–4^, COVID-19 patients presented with marked reductions in T cell number and frequency in both CD4^+^ and CD8^+^ T cells, even after normalization for age as a possible confounder (Extended Data Fig.1d). Granulocytes such as neutrophils and eosinophils are normally excluded from the PBMC fraction following density gradient separation. However, low density granulocytes are present in the PBMC layer from peripheral blood collections in patients with inflammatory diseases^17^. We observed an increases in monocytes, and low density neutrophils and eosinophils that correlated with the severity of disease (Fig. 2c, Extended Data Fig. 2a,b). Additionally, we observed increased activation of T cells and a reduction in HLA-DR expression by circulating monocytes^1^ (Extended Data Fig. 2c). A complete overview of PBMC cells subsets is presented in Extended Data Fig. 2.

**Figure 2:**
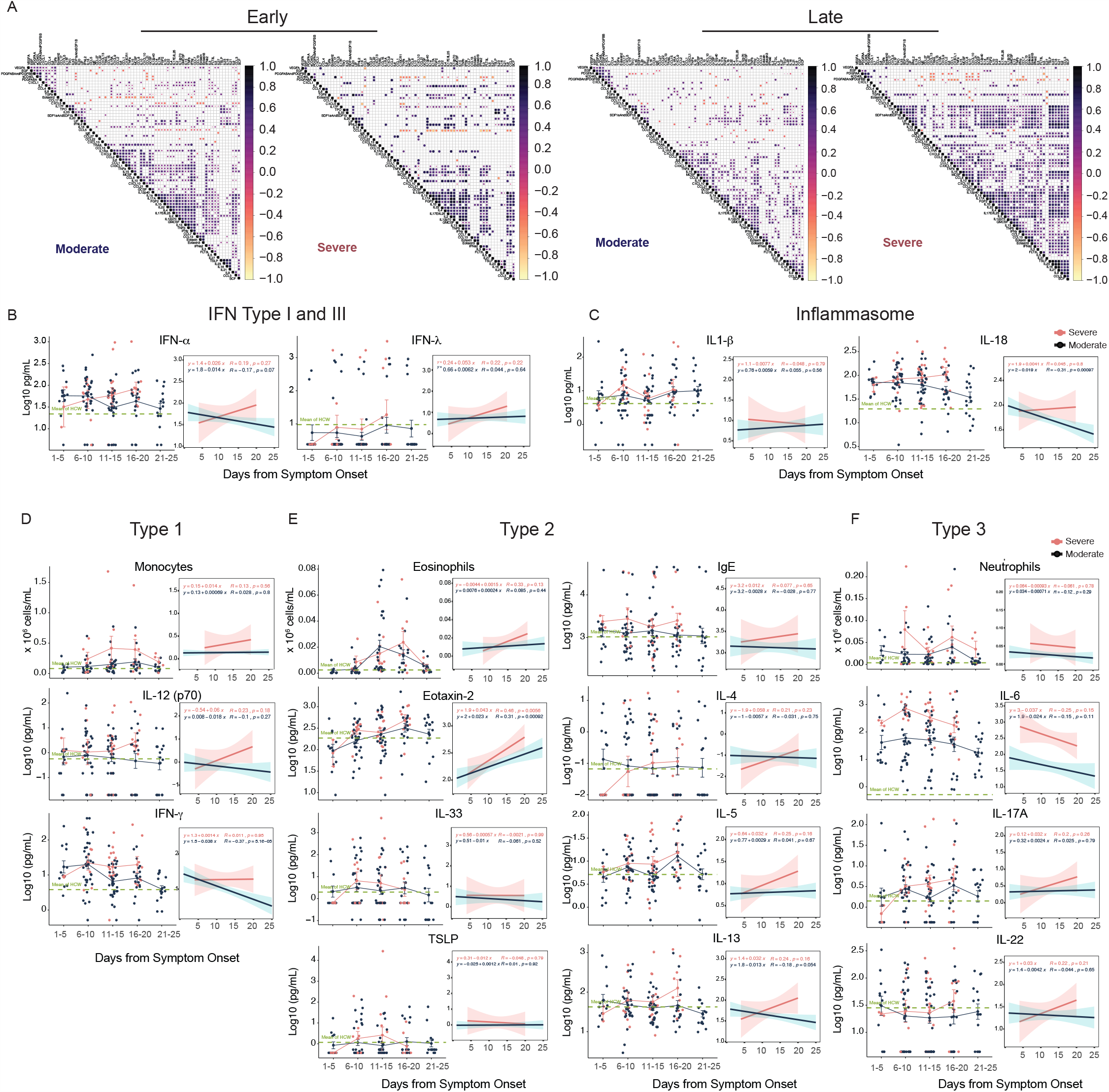
Longitudinal immune profiling of moderate and severe COVID-19 patients. **a**, Correlation matrices of 71 cytokines from patient blood comparing cytokine concentrations within moderate patients or severe patients during the early phase (less than 10 days following symptom onset) or late phase (greater than 10 days following symptom onset) of disease. Only significant correlations are represented as dots, and Pearson’s correlation coefficient from comparisons of cytokine measurements within each patient is visualized by colour intensity. **b**, Anti-viral Interferons, **(c)** inflammasome-related cytokines measured as Log10 concentration and plotted over time according to the days of symptom onset for patients with moderate disease (n=112) or severe disease (n=39). Each dot represents a distinct patient and time point arranged by intervals of five days until 25 days. The dotted green line represents the mean measurement from uninfected health care workers. Longitudinal data was also plotted over time continuously according to days following symptom onset. Regression lines are indicated by the dark blue (moderate) or red (severe) solid lines. Associated linear regression equations, Pearson’s Correlation Coefficients, and significance are in pink (moderate) or dark blue (severe). 95% confidence intervals for the regression lines are denoted by the pink (moderate) or dark blue (severe) filled areas. **d-f**, Cellular and cytokine measurements representative of **(d)** Type1, **(e**) Type2, **(f)** Type3 immune responses reported over time in intervals of days or continuously as linear regressions.

Severe disease caused by infections with MERS, SARS-CoV-1, and SARS-CoV-2 coronaviruses are associated with cytokine release syndrome (CRS)^1,2,8,11,12,18,19^. To gain insights into key differences in cytokines, chemokines, and additional immune markers between moderate and severe patients, we correlated the measurements of these soluble proteins across all patients’ time-points that were collected (Fig. 1d). We observed a “core COVID-19 signature” shared by both moderate and severe groups of patients defined by the following inflammatory cytokines that positively correlated with each other; these include:IL-1α, IL-1β, IL-17A, IL-12 p70, and IFN-α (Fig. 1d). In severe patients, we observed an additional inflammatory cluster defined by: TPO, IL-33, IL-16, IL-21, IL-23, IFN-λ, eotaxin and eotaxin 3 (Fig. 1d). Most of the cytokines linked to CRS, such as IL-1α, IL-1β, IL-6, IL-10, IL-18 and TNF-α, showed increased positive associations in severe patients (Fig. 1d, e, f and Extended Data Fig. S3). These data highlight the broad inflammatory changes, involving concomitant release of type-1, type-2 and type-3 cytokines in severe COVID-19 patients.

### Longitudinal immune profiling of moderate and severe COVID-19 patients

Our data presented above, as well as previous single-cell transcriptome and flow cytometry-based studies ^2,4,19–21^, depicted an overt innate and adaptive immune activation in severe COVID-19 patients. We sought to determine the temporal dynamics of immune changes through longitudinal sample collection and analysis of key immunological markers in moderate versus severe patients. Longitudinal cytokines correlations, measured as days from symptom onset (DfSO), indicated major differences in immune phenotype between moderate and severe disease apparent after day 10 of infection (Fig. 2a). In the first 10 DfSo, severe and moderate patients displayed similar correlation intensity and markers, including the overall “core COVID-19 signature” described above (Fig. 2a). However, after day 10 these markers steadily declined in patients with moderate disease; in contrast, severe patients maintained elevated levels of these core signature makers. Notably, additional correlations between cytokines emerged in patients with severe disease following day 10 (Fig. 2a). These analyses strongly support the observation in the overall analysis described in Fig. 1, in which TPO and IFN-α strongly associated with IFN-λ, IL-9, IL-18, IL-21, IL-23, and IL-33 (Fig. 2a). These observations indicate sharp differences in the expression of inflammatory markers along disease progression between patients who exhibit moderate vs. severe COVID-19 symptoms.

Temporal analyses of PBMC and soluble proteins in plasma, either by linear regression or grouped intervals, supported distinct courses in disease. IFN-α levels were sustained at higher levels in sever patients while they declined moderate patients (Fig. 2b). Plasma IFN-λ levels increased during the first week of symptom onset in ICU patients and remained elevated in later phases (Fig. 2b). Additionally, inflammasome-induced cytokines, such as IL-1β and IL-18 were also elevated in severe patients compared to patients with moderate disease at most time-points analysed (Fig. 2c). Consistently, IL-1 receptor antagonist (IL-1Ra), induced by IL-1R signalling as a negative feedback regulator^22^, also showed increased levels in ICU patients from day 10 of disease onset (Extended Data Fig. 4).

With respect to type-1 immunity, an increased number of monocytes was observed at approximately 14 DfSo in severe but not in moderate COVID-19 patients (Fig. 2d). The innate cytokine IL-12, a key inducer of type-1 immunity^13,14^, displayed a similar pattern to IFN-γ; increasing over time in severe patients but steadily declining in moderate patients (Fig. 2d). IFN-γ can be secreted by ILC1, NK, and Th1 cells. By intracellular cytokine staining, CD4^+^ and CD8^+^ T cells from patients with moderate disease secreted comparable amounts of IFN-γ to those from severe patients. Together with the severe T cell depletion in severe patients (Fig. 1), our data suggested that secretion of IFN-γ by non-T cells (ILC1, NK), or non-circulating T cells in tissues were the primary contributors to the enhanced levels observed in severe patients (Extended Data Fig. 5).

Type-2 immune markers continued to increase in severe patients over time, as indicated by strong correlations observed in late time points from severe patients (Fig. 2a). Eosinophils and eotaxin-2 increased in severe patients and remained higher than levels measured in moderate patients (Fig. 2e). Type-2 innate immune cytokines, including TSLP and IL-33, did not exhibit significant differences between severe and moderate patients (Fig. 2e). Hallmark type-2 cytokines, including IL-5 (associated with eosinophilia) and IL-13 (Fig. 2e), were enhanced in patients with severe over moderate disease. In contrast, IL-4, was not significantly different. However, IL-4, similar to IL-5 and IL-13, exhibited an upward trend over the course of disease in severe patients (Fig. 2e). A type-2 antibody isotype was also increased; IgE levels were significantly higher in severe patients and continued to increase during the disease course (Fig. 2e).

IL-6 linked to CRS was significantly elevated in severe patients, although circulating neutrophils did not show a significant increase in our longitudinal analysis (Fig. 2f). Hallmarks of type-3 responses were observed in severe patients, including increased plasma IL-17A and IL-22, as well as IL-17 secretion by circulating CD4 T cells as assessed by intracellular cytokine staining (Fig. 2f, Extended Data Fig. 5). These data identify broad elevation of type-1, type-2 and type-3 signatures in severe cases of COVID-19, with distinct temporal dynamics and quantities between severe and moderate patients.

### Nasopharyngeal viral load correlates with elevated cytokines

We next asked which early immunological profile was correlated with worse disease trajectory and whether these parameters were influenced by the viral load. We first measured viral load kinetics by serial nasopharyngeal swabs. While viral RNA load was not significantly different at any specific time point analysed post symptom onset between severe and moderate patients, moderate patients showed a steady decline in viral load over the course of disease, and severe patients did not (Fig. 3a). Regardless of whether the patients exhibited moderate or severe disease, viral load significantly correlated with the levels of IFN-α, IFN-γ, TNF-α and TRAIL (Fig. 3b). Additionally, several chemokines responsible for monocyte recruitment significantly correlated with viral load only in patients with severe disease (Extended Data Fig. 6a,b). These data indicated that nasopharyngeal viral load positively correlates with plasma levels of interferons and cytokines.

**Figure 3.**
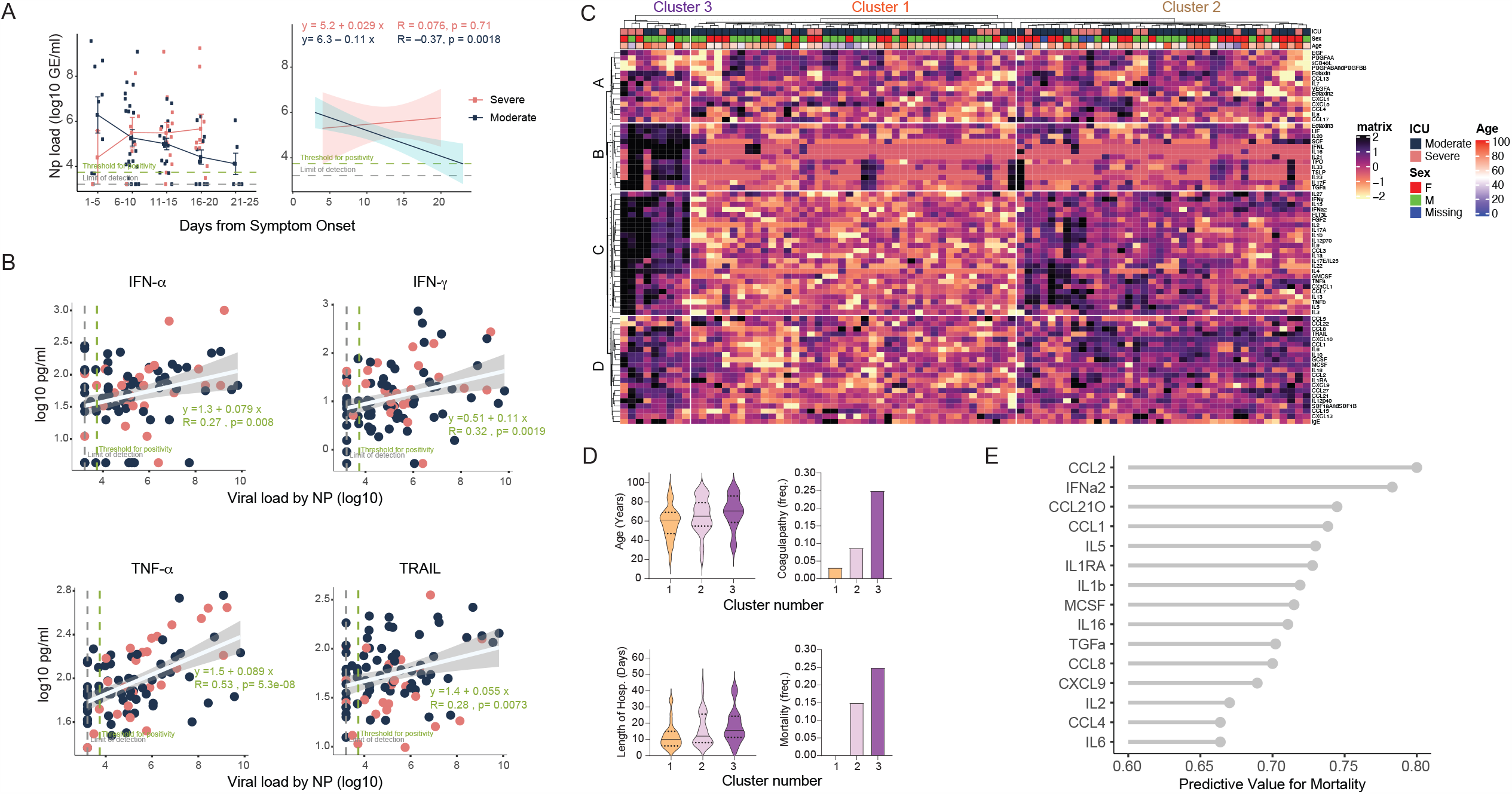
Early viral and cytokine profiles distinguish moderate and severe outcomes. **a**, Viral loads measured by nasopharyngeal swabs were plotted as Log10 of genome equivalents time according to the days following symptom onset for patients with moderate disease (n=112) or severe disease (n=39). Each dot represents a distinct patient and time point arranged by intervals of five days until 25 days. Longitudinal data was also plotted over time continuously according to days following symptom onset. Regression lines are indicated by the dark blue (moderate) or red (severe) solid lines. Associated linear regression equations, Pearson’s Correlation Coefficients, and significance are in pink (moderate) or dark blue (severe). Text in green denotes the regression analysis and correlation for all patients. 95% confidence intervals for the regression lines are denoted by the pink (moderate) or dark blue (severe) filled areas. **b**, Correlation and linear regression of cytokines plotted, as Log10 of concentration, and viral load by nasopharyngeal swab, plotted as Log10 of genome equivalents, regardless of disease severity (n=151). Each dot represents a unique patient time point classified as experiencing either moderate (dark blue) or severe (red) disease. The regression line for all patients is indicated by the white solid line. The associated linear regression equation, Pearson’s Correlation Coefficient, and significance is denoted in green text. The 95% confidence intervals for the regression line is indicated by the grey filled areas. **c**, Unbiased heat map comparisons of cytokines within peripheral blood mononuclear cells (PBMCs) measured until 12 days after symptom onset in COVID-19 patients. Measurements were normalized across all patients. K-means clustering was used to determine Clusters 1-3 (Cluster 1, n=46; Cluster 2, n=50; Cluster 3, n=16). **d**, Age, coagulopathy, length of hospitalization and mortality within each cluster, previously determined. **e**, The top 20 cytokines by mutual information analysis to determine their respective importance for determining mortality. Significance of comparisons were determined by Wilcoxon Rank-Sum Test and indicated as such: * p < 0.05, ** p< 0.01, *** p < 0.001, and **** p <0.0001.

**Figure 4.**
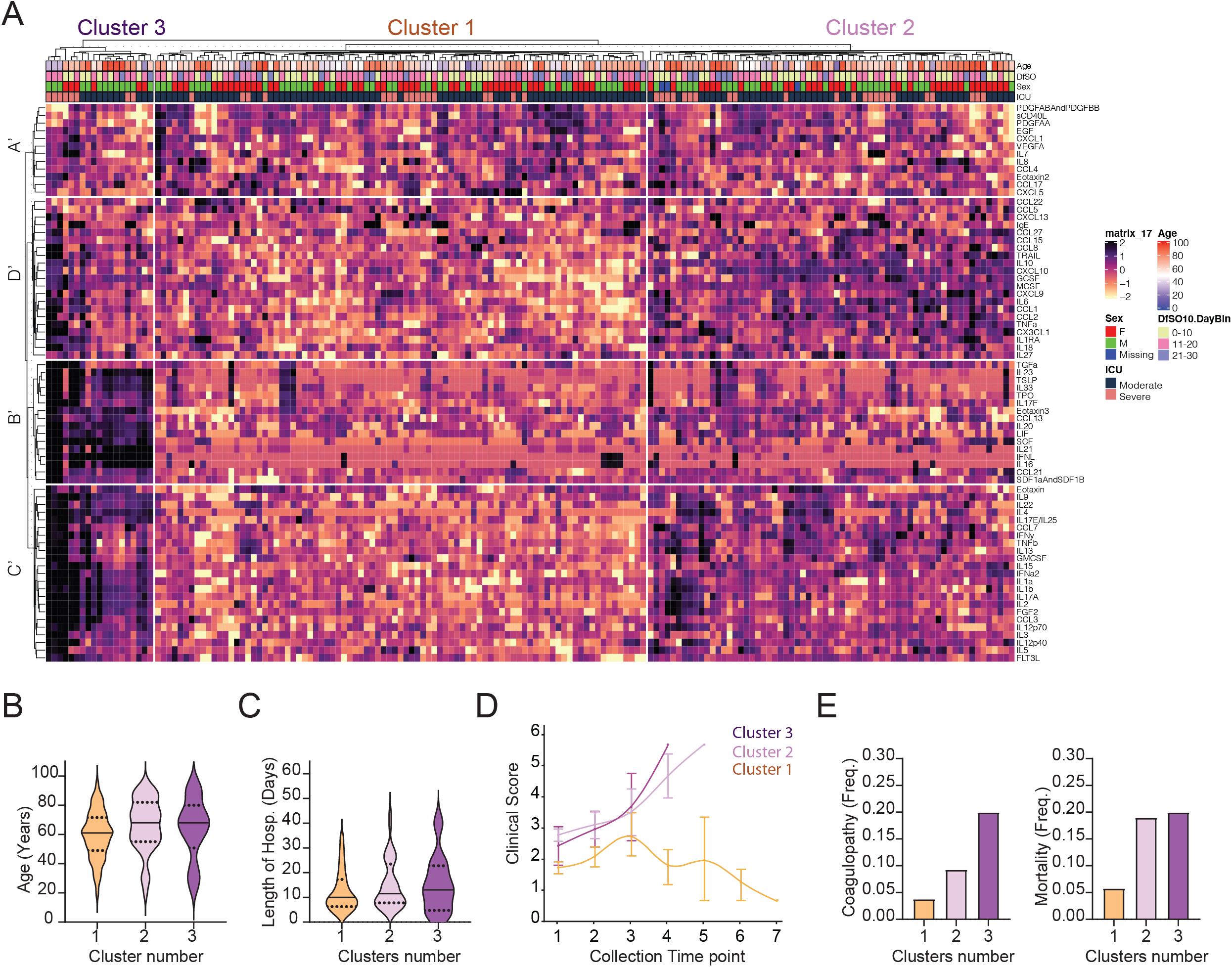
Immune correlates of COVID-19 outcomes. **a**, Unbiased heat map comparisons of cytokines within peripheral blood mononuclear cells (PBMCs) measured at distinct time points in COVID-19 patients. Measurements were normalized across all patients. K-means clustering was used to determine Clusters 1-3 (Cluster 1, n=84; Cluster 2, n=66; Cluster 3, n=20). **b**, Age distribution and length of hospitalization of patients within each cluster, previously determined. Solid lines indicate the mean measurement; dotted lines indicate the standard deviation. **c**, Proportion of patients of male or female within each cluster. **d**, Disease progression, according to the assigned clinical severity scale, for patients belonging to each cluster. Time was ordered by the collection time points for each patient, with regular collection intervals of 3-4 days as indicated on **Extended data fig**.**7. e**, Proportion of patients in each cluster with coagulopathies or ultimately fatal outcome.

### Early cytokine profile distinguishes moderate and severe disease outcomes

Next, we examined whether specific early cytokine responses are associated with severe COVID-19. To this end, we conducted an unsupervised clustering analysis using patients’ baseline measurements, collected before 12 DfSO (Fig. 3c). Three main clusters with correlation to distinct disease outcome emerged. These were characterized by 4 distinct immune signatures: Signature A contained several stromal growth factors including EGF, PDGF, VEGF that are mediators of wound healing and tissue repair^23^, as well as IL-7, a critical growth factor for lymphocytes. Signature B consisted of eotaxin 3, IL-33, TSLP, along with IL-21, IL-23 and IL-17F, thus representing type-2 & 3 immune effectors. Signature C comprised of mixtures of all immunotypes, including type-1 cytokines (IFN-γ, IL-12 p70, IL-15, IL-2, TNF-α), type-2 (IL-4, IL-5, IL-13), as well as type-3 (IL-1α, IL-1β, IL-17A, IL-17E, IL-22). Finally, signature D contained a number of chemokines involved in leukocyte trafficking including CCL1, 2, 5, 8, 15, 21, 22, 27, CXCL9, 10, 13, and SDF1.

Cluster 1 was comprised primarily of patients with moderate disease who experienced low occurrences of coagulopathy, shorter lengths of hospital stay, and no mortality (Fig. 3c, d). The main characteristics in this cluster were low levels of inflammatory markers and similar or increased levels of parameters in signature “A” containing tissue reparative growth factors (Fig. 3c). Clusters 2 and 3 were characterized by the rise in inflammatory markers, and patients belonging to these clusters had higher incidence of coagulopathy and mortality, which was more pronounced in cluster 3 (Fig. 3c,d). Cluster 2 showed higher levels of markers in signatures “C and D”, which included IFN-α, IL-1Ra and several hallmark type-1, type-2 and type-3 cytokines, than patients in cluster 1, but lower expression of markers in signatures “B, C and D” than in Cluster 3 (Fig. 3c,d). Cluster 3 displayed heightened expression of markers in signatures “B, C and D” than other clusters. Cluster 3 showed particular enrichment in expression of markers in signature “B”, which include several innate cytokines including IFN-λ, TGF-α, TSLP, IL-16, IL-23 and IL-33, and markers linked to coagulopathy, such as TPO (Fig. 3c, d).

We next ranked these parameters obtained at early time points as predictors of severe disease outcomes (Fig. 3e, Extended Data Fig. 6c). In both cases, plasma inflammatory markers strongly associated with severe disease outcomes. For example, high levels of type I IFN (IFN-α) before the first 12 DfSo correlated with longer hospitalization time and death (Fig. 3e, Extended Data Fig. 6c). Moreover, patients who ultimately died of COVID-19 exhibited significantly elevated levels of IFN-α, IFN-λ, IL-1Ra as well as chemokines associated with monocytes and T cells recruitment and survival, including CCL1, CLL2, MCSF, IL-2, IL16 and CCL21 before the first 12 DfSO (Fig. 3e, Extended Data Fig. 6c). These analyses identify specific immunological markers that appear early in the disease that strongly correlate with worse outcomes and death.

### Retrospective analysis of immune correlates of COVID-19 outcomes

To further evaluate potential drivers of severe COVID-19 outcome in an unbiased manner, we performed unsupervised clustering analysis including all patients and all timepoints using cytokines and chemokines (Fig. 4a). Notably, three main clusters of patients emerged and the distribution of patients in early time-point clusters identified in Fig. 3c matched the distribution for the all-time point analysis (Fig. 4a) in 96% of cases. Cluster 1 was comprised primarily of moderate disease patients and showing improving clinical signs (Fig. 4a-d, Extended Data Fig. 7). This cluster contained only two deceased patients. Cluster 1 was characterized by low levels of inflammatory markers as well as similar or increased expression of markers in the signature A’ (Fig. 4a-d), which mostly matched signature A markers described in Fig. 3c. Clusters 2 and 3 contained patients displaying coagulopathy and worsened clinical progression, including higher number of deceased patients (Fig. 4a-d, Extended Data Fig. 7).

Clusters 2 and 3 were driven by a set of inflammatory markers falling into signatures B’, C’ and D’ to some extent, which highly overlap with the “core signature” cytokines and chemokines identified in Fig. 1 as well as the signatures “B and C” identified in Fig. 3c. These include type-1 immunity markers, including IL-12, chemokines linked to monocyte recruitment and IFN-γ, type-2 responses, such as TSLP, chemokines linked to eosinophil recruitment, IL-4, IL-5 and IL-13, and type-3 responses, including IL-23, IL-17A and IL-22. Additionally, most CRS and inflammasome-associated cytokines were enriched in these clusters, including IL-1α, IL-1β, IL-6, IL-18 and TNF-α (Fig. 4a). These findings were consistent with generalized estimating equations that identified relationships between biomarkers death over time (Extended Data Fig. 8). Together, these results identify groups of inflammatory, as well as potentially protective, markers that correlated with COVID-19 trajectory. The immune signatures that correlate with recovery (cluster 1) and worsening diseases (cluster 2 < cluster 3) were remarkably similar whether we took prospective (Fig. 3) vs. retrospective (Fig. 4) approaches.

## Discussion

Our longitudinal analyses of hospitalized COVID-19 patients revealed key temporal features of viral load and immune responses that distinguish disease trajectories during hospitalization. Using patients’ immune response readouts, unsupervised clustering revealed 3 distinct profiles that influence the evolution and severity of COVID-19. Cluster 1, characterized by low expression of proinflammatory cytokines and enrichment in tissue repair genes, followed a disease trajectory that remained moderate leading to eventual recovery. Clusters 2 and 3 were characterized with highly elevated proinflammatory cytokines (cluster 3 being more intense), developed worse disease and many died of COVID-19. Thus, in addition to the well-appreciated CRS-related pro-inflammatory cytokines, we propose four signatures of immune response profiles that more accurately subset patients into distinct COVID-19 disease course.

In addition to dynamic changes in leukocyte subsets, such as prolonged lymphopenia and elevation of monocytes and neutrophils in severe disease, we observed an increase in eosinophils in patients admitted to the ICU. By focusing on hallmark cytokines of type 1, type 2 and type 3 immunity, we found that while all are higher in severe disease than moderate disease, type 2 signatures, including eotaxin 2, IgE, eosinophils, IL-5 and IL-13, continue to become elevated during disease course in the severe patients. While nasopharyngeal viral RNA levels were not significantly different between moderate and severe patients at the specific time points examine post symptom onset, linear regression analyses showed slower decline in viral load in patients admitted to the ICU. Viral load was highly correlated with IFN-α, IFN-γ and TNF-α, suggesting that viral load may drive these cytokines, and that interferons do not successfully control the virus. Moreover, many interferons, cytokines, and chemokines were elevated early in disease for patients who ultimately died of COVID-19. This suggests possible pathological roles associated with these host defence factors.

Our comprehensive analysis of soluble plasma factors revealed a broad misfiring of immune effectors in COVID-19 patients, with early predictive markers, distinct dynamics between types of immune responses, among moderate and severe disease outcomes. Unsupervised clustering identified a set of inflammatory markers highly enriched in severe patients. By following these patients over time (Figure 2), we observed a mixed response of type-1, −2 and −3-associated cytokines. A preprint study assessing inflammatory cytokines increased in COVID-19 in comparison to influenza patients only observed heightened IL-6 and IL-8 (Ref. ^24^); the latter was not enriched in clusters 2 or 3 in our analysis. Interestingly, this study also observed high circulating levels of IL-1Ra in severe patients, which we also identified. We likewise observed that IL-6 may be an important clinical target in COVID-19. It was both highly enriched in patients with severe disease in clusters 2 and 3. In fact, all of our ICU patients, including the ones who succumbed to the disease, received Tocilizumab, an IL-6R blocking antibody, according to a protocol uniquely instituted at <u>Yale New Haven Hospital COVID-19</u>. Recent studies reported positive outcomes with this treatment, including a reduction in an inflammatory-monocyte population associated with worse outcomes^25^. This highlights the need for combination therapy to block other cytokines highly represented by these clusters, including inflammasome-dependent cytokines and type-2 cytokines.

Our data, suggest a pathological role for type-2 immune responses to COVID-19 (Ref. ^26^). Previous studies have proposed a pathogenic role for type-2 immunity in influenza, as well as SARS-CoV infection^27,28^. In addition to key cytokines associated with type-2 immunity, including IL-5 and IL-13, we observed an early and persistent increase in IgE levels in severe COVID-19 patients. Interestingly, we also observed transient increases in circulating eosinophil levels - paralleled by simultaneous elevations in Type II cytokines - among severe COVID-19 patients despite widespread use of glucocorticoids among this population (Extended Data Fig. 1c). Given the potent eosinopenia induced by glucocorticoids, further study exploring the extent of margination of Type II effector cells into COVID-19 affected tissues, and their role in disease progression, is warranted^29^. A retrospective cohort study, also deposited as preprint, reported positive clinical outcomes upon administration of famotidine, a histamine antagonist, to COVID-19 patients^30^. Similarly, a preprint publication analysing regulatory networks downstream of ACE2 and the spike protein activator, TMPRSS2, which is part of the mucus secretory pathway, speculated that upregulation of type-2 immunity and IFN-responses, could increase ACE2 expression thus aggravating disease^31,32^.

Another notable observation from this study is the overwhelming association between antiviral IFNs and disease. While type I and III IFNs are generally required for viral clearance, during persistent viral infection, or under high viral loads, conditions generally observed in elderly patients, IFN responses can lead to pathological outcomes^33^. Our study found that IFN-α is elevated in patients with severe disease, and higher levels correlate with death. The correlation between increased viral load and increased secretion of type I IFN suggest that persistence of virus drives type I IFN secretion, but the prolonged secretion and failure to control virus instead leads to a proinflammatory cytokine storm. This vicious cycle can lead to the worsening of disease symptoms, ventilator use and ultimately death.

In addition to CRS-associated cytokines, we observed a correlation with cytokines linked to the inflammasome pathway, which partially overlap with CRS, including IL-1β and IL-18. Indeed, it is plausible that inflammasome activation, along with a sepsis-like CRS, triggers vascular insults or tissue pathology observed in severe COVID-19 patients ^34^. The immune dysfunction observed in severe COVID-19 patients included hallmarks of type-3 cytokines, IL-17 and IL-22. The primary function of IL-17 function is tissue neutrophil recruitment, which is associated with COVID-19 severity. Additionally, IL-17 is known to enhance the production of inflammatory cytokines associated with COVID-19, as well as lung pathology, including TNF-α, IL-1β and IL-6 ^35^. Our results suggest that a multi-faceted inflammatory response is associated with late COVID-19 severity. This raises the possibility that early immunological interventions that target inflammatory markers predictive of worse disease outcome are preferred to blocking late-appearing cytokines. Overall, our analyses provide a comprehensive examination of the diverse inflammatory dynamics during COVID-19 and possible contributions by distinct sets of inflammatory mediators towards disease progression. Our disease trajectory analyses provide bases for more targeted treatment of COVID-19 patients based on early cytokine markers, as well as therapies targeted to enhancing tissue repair and promoting disease tolerance.

## Data Availability

The data that support the findings of this study will be made openly available in public repositories.

## Methods

### Ethics statement

This study was approved by Yale Human Research Protection Program Institutional Review Boards (FWA00002571, Protocol ID. 2000027690). Informed consents were obtained from all enrolled patients and healthcare workers.

### Patients

135 COVID-19 patients admitted to Yale-New Haven Hospital (YNHH) between March 18th and May 5th, 2020 were included in this study. Nasopharyngeal swabs were collected, as recently described^36^, approximately every four days for SARS-CoV-2 RT-qPCR analysis where clinically feasible. Paired whole blood for flow cytometry analysis was collected simultaneously in sodium heparin-coated vacutainers and kept on gentle agitation until processing. All blood was processed the same day as collection from patients. Patients were scored for COVID-19 disease severity through review of electronic medical records (EMR) at each longitudinal time point. Scores were assigned by a clinical infectious disease physician according to a custom developed disease severity scale. Moderate disease status (Clinical Score 1, 2 and 3) was defined as: (1) SARS-CoV-2 infection requiring hospitalization without supplemental oxygen, (2) infection requiring non-invasive supplemental oxygen (<3 L / min, sufficient to maintain greater than 92% SpO_2_), (3) infection requiring non-invasive supplemental oxygen (> 3L supplemental oxygen to maintain SpO2 > 92%, or, required > 2L supplemental oxygen to maintain SpO2 > 92% and had a high sensitivity C-reactive protein (CRP) > 70) and received tocilizumab. Severe disease status (Clinical score 4 and 5) was defined as infection meeting all criteria for clinical score 3 while also requiring admission to the YNHH Intensive Care Unit (ICU) and > 6L supplemental oxygen to maintain SpO2 > 92% (4); or infection requiring invasive mechanical ventilation / extracorporeal membrane oxygenation (ECMO) in addition to glucocorticoid / vasopressor administration (5). Clinical score 6 was assigned for deceased patients. The use of tocilizumab can increase circulating levels of IL-6 through inhibition of IL-6Rα-mediated degradation. Analysis of our cohort indicate higher plasma levels of IL-6 in both moderate and severe patients that received tocilizumab treatment (Extended data Fig. 1d).

For all patients, days from symptom onset were estimated according to the following scheme: (1) highest priority was given explicit onset dates provided by patients; (2) next highest priority was given to the earliest reported symptom by a patient, and (3) in the absence of direct information regarding symptom onset, we estimated a date through manual assessment of the electronic medical record (EMRs) by an independent clinician. Demographic information was aggregated through a systematic and retrospective review of patient EMRs and was used to construct Extended Table 1. Symptom onset and etiology was recorded through standardized interview with patients or patient surrogates upon enrollment in our study, or alternatively through manual EMR review if no interview was possible due to clinical status.

### Viral RNA measurements

RNA concentrations were measured from nasopharyngeal samples by RT-qPCR as previously described^36^. Briefly, total nucleic acid was extracted from 300 μl of viral transport media (nasopharyngeal swab) using the MagMAX Viral/Pathogen Nucleic Acid Isolation kit (ThermoFisher Scientific) using a modified protocol and eluted into 75 μl of elution buffer.

For SARS-CoV-2 RNA detection, 5 μl of RNA 371 template was tested as previously described^37^, using the US CDC real-time RT-qPCR primer/probe sets for 2019-nCoV_N1, 2019-nCoV_N2, and the human RNase P (RP) as an extraction control. Virus RNA copies were quantified using a 10-fold dilution standard curve of RNA transcripts that we previously generated^37^. The lower limit of detection for SARS-CoV-2 genomes assayed by qPCR in nasopharyngeal specimens was established as recently described^37^. In addition to a technical detection threshold, we also utilized a clinical referral threshold (detection limit) to either: (1) refer asymptomatic HCWs for diagnostic testing at a CLIA-approved laboratory, or (2) cross-validate results from a CLIA-approved laboratory for SARS-CoV-2 qPCR+ individuals upon study enrollment. Individuals above the technical detection threshold, but below the clinical referral threshold, are considered SARS-CoV-2 positive for the purposes of our research study.

### Isolation of patient plasma

Plasma samples were collected after whole blood centrifugation at 400 g for 10 minutes at RT without brake. The undiluted serum was then transferred to 15 ml polypropylene conical tubes, and aliquoted and stored at −80 °C for subsequent analysis.

### Cytokine and chemokine measurements

Patient serum was isolated as before and aliquots were stored in −80°C. Sera were shipped to Eve Technologies (Calgary, Alberta, Canada) on dry ice, and levels of cytokines and chemokines were measured with Human Cytokine Array/Chemokine Array 71-403 Plex Panel (HD71). All the samples were measured upon the first thaw.

### Isolation of PBMCs

Peripheral blood mononuclear cells (PBMCs) were isolated from heparinized whole blood using Histopaque (Sigma-Aldrich, #10771-500ML) density gradient centrifugation in a biosafety level 2+ facility. After isolation of undiluted serum, blood was 1:1 diluted in room temperature PBS and layered over Histopaque in a SepMate tube (Stemcell Technologies; #85460) and centrifuged for 10 minutes at 1200g. The PBMC layer was isolated according to manufacturer’s instructions. Cells were washed twice with PBS prior to counting. Pelleted cells were briefly treated with ACK lysis buffer for 2 minutes and then counted. Percentage viability was estimated using standard Trypan blue staining and an automated cell counter (Thermo-Fisher, #AMQAX1000).

### Flow cytometry

Antibody clones and vendors are as follows: BB515 anti-HLA-DR (G46-6), BV785 anti-CD16 (3G8), PE-Cy7 anti-CD14 (HCD14), BV605 anti-CD3 (UCHT1), BV711 anti-CD19 (SJ25C1), BV421 anti-CD11c (3.9), AlexaFluor647 anti-CD1c (L161), Biotin anti-CD141 (M80), PE anti-CD304 (12C2), APCFire750 anti-CD11b (ICRF44), PerCP/Cy5.5 anti-CD66b (G10F5), BV785 anti-CD4 (SK3), APCFire750 or PE-Cy7 or BV711 anti-CD8 (SK1), BV421 anti-CCR7 (G043H7), AlexaFluor 700 anti-CD45RA (HI100), PE anti-PD1 (EH12.2H7), APC anti-TIM3 (F38-2E2), BV711 anti-CD38 (HIT2), BB700 anti-CXCR5 (RF8B2), PE-Cy7 anti-CD127 (HIL-7R-M21), PE-CF594 anti-CD25 (BC96), BV711 anti-CD127 (HIL-7R-M21), BV421 anti-IL17a (N49-653), AlexaFluor 700 anti-TNFa (MAb11), PE or APC/Fire750 anti-IFNy (4S.B3), FITC anti-GranzymeB (GB11), AlexaFluor 647 anti-IL4 (8D4-8), BB700 anti-CD183/CXCR3 (1C6/CXCR3), PE-Cy7 anti-IL-6 (MQ2-13A5), PE anti-IL-2 (5344.111), BV785 anti-CD19 (SJ25C1), BV421 anti-CD138 (MI15), AlexaFluor700 anti-CD20 (2H7), AlexaFluor 647 anti-CD27 (M-T271), PE/Dazzle594 anti-IgD (IA6-2), PE-Cy7 anti-CD86 (IT2.2), APC/Fire750 anti-IgM (MHM-88), BV605 anti-CD24 (M1/69), APC/Fire 750 anti-CD10 (HI10a), BV421 anti-CD15 (SSEA-1), AlexaFluor 700 Streptavidin (ThermoFisher). Briefly, freshly isolated PBMCs were plated at 1-2 x 10^6^ cells / well in a 96 well U-bottom plate. Cells were resuspended in Live/Dead Fixable Aqua (ThermoFisher) for 20 minutes at 4°C. Following a wash, cells were then blocked with Human TruStan FcX (BioLegend) for 10 minutes at RT. Cocktails of desired staining antibodies were directly added to this mixture for 30 minutes at RT. For secondary stains, cells were first washed and supernatant aspirated; then to each cell pellet a cocktail of secondary markers was added for 30 minutes at 4°C. Prior to analysis, cells were washed and resuspended in 100 μL of 4% PFA for 30 minutes at 4°C. For intracellular cytokine staining following stimulation, cells were resuspended in 200 μL cRPMI (RPMI-1640 supplemented with 10% FBS, 2 mM L-glutamine, 100 U/ml penicillin, and 100 mg/ml streptomycin, 1mM Sodium Pyruvate, and 50uM 2-Mercaptoethanol) and stored at 4°C overnight. Subsequently, these cells were washed and stimulated with 1X Cell Stimulation Cocktail (eBioscience) in 200 μL cRPMI for 1 hour at 37°C. 50 μL of 5X Stimulation Cocktail (plus protein transport 442 inhibitor) (eBioscience) was added for an additional 4 hours of incubation at 37°C. Following stimulation, cells were washed and resuspended in 100 μL of 4% PFA for 30min at 4°C. To quantify intracellular cytokines, these samples were permeabilized with 1X Permeabilization Buffer from the FOXP3/ Transcription Factor Staining Buffer Set (eBioscience) for 10 minutes at 4°C. All subsequent staining cocktails were made in this buffer. Permeabilized cells were then washed and resuspended in a cocktail containing Human TruStan FcX (BioLegend) for 10 minutes at 4°C. Finally, intracellular staining cocktails were directly added to each sample for 1 hour at 4°C. Following this incubation, cells were washed and prepared for analysis on an Attune NXT (ThermoFisher). Data were analysed using FlowJo software version 10.6 software (Tree Star). The specific et of markers used to identify each subset of cells are summarized in Extended Data Table 8.

### Statistical analysis

The patients and its measurements were clustered using the kmeans algorithm available within the ComplexHeatmap package^38^. To determine the optimum number of clusters, we ran the data matrix within the NBClust package^39^. Prior to data visualization, each patient was scaled by subtracting each measurement with the mean and dividing by the standard deviation. Multiple group comparisons were analyzed by running both parametric (ANOVA) and non-parametric (Kruskal-Wallis) statistical tests with the Dunn’s Test post hoc tests. Mutual information analyses were performed using the Caret R package and visualized with ggPlot2. Multiple correlation analysis was performed by executing spearman correlations with the Hmisc R package and visualized with corrplot, only showing correlations with pvalue less than 0.05. For generalized linear models (GLM), we calculated the incident risk ratio (IRR) by conducting a Poisson regression with a log link and robust variance estimation; this value approximates the risk ratio estimated by a log-linear model. For generalized estimating equation models (GEE), we calculated the incidence risk ratio (IRR) in the same way as for non-GEE GLM models, assuming an independent correlation structure. All models controlled for participant sex and age.

### Yale IMPACT Research Team Authors

Abeer Obaid, Alice Lu-Culligan, Allison Nelson, Anderson Brito, Angela Nunez, Anjelica Martin, Annie Watkins, Bertie Geng, Chaney Kalinich, Christina Harden, Codruta Todeasa, Cole Jensen, Daniel Kim, David McDonald, Denise Shepard, Edward Courchaine, Elizabeth B. White, Erin Silva, Eriko Kudo, Giuseppe DeIuliis, Harold Rahming, Hong-Jai Park, Irene Matos, Jessica Nouws, Jordan Valdez, Joseph Fauver, Joseph Lim, Kadi-Ann Rose, Kelly Anastasio, Kristina Brower, Laura Glick, Lokesh Sharma, Lorenzo Sewanan, Lynda Knaggs, Maksym Minasyan, Maria Batsu, Mary Petrone, Maxine Kuang, Maura Nakahata, Melissa Campbell, Melissa Linehan, Michael H. Askenase, Michael Simonov, Mikhail Smolgovsky, Nicole Sonnert, Nida Naushad, Pavithra Vijayakumar, Rick Martinello, Rupak Datta, Ryan Handoko, Santos Bermejo, Sarah Prophet, Sean Bickerton, Sofia Velazquez, Tara Alpert, Tyler Rice, William Khoury-Hanold, Xiaohua Peng, Yexin Yang, Yiyun Cao, Yvette Strong.

## Acknowledgements

We thank Melissa Linehan for technical and logistical assistance, and thank helpful discussions with Drs. Andrew Wang, Aaron Ring, Craig Wilen and Daniel Mucida. This work was supported by the Women’s Health Research at Yale Pilot Project Program (AI, AR), Fast Grant from Emergent Ventures at the Mercatus Center, Mathers Foundation, and the Ludwig Family Foundation, the Department of Internal Medicine at the Yale School of Medicine, Yale School of Public Health and Beatrice Kleinberg Neuwirth Fund. A.I. is an Investigator of the Howard Hughes Medical Institute. C.L. is a Pew Latin American Fellow. P.Y is supported by Gruber Foundation and the NSF. B.I is supported by NIAID 2T32AI007517-16. CBFV is supported by NOW Rubicon 019.181EN.004.

## Author Contributions

C.L. and A.I. conceived the study. A.I.K. led the IMPACT biorepository. C.L., P.W., J.K., designed the experiments, defined parameters, collected and processed patient PBMC samples and analyzed data. J.S., J.E.O., T.M., S.M., H.W. processed PBMC samples. T.B.R.C. performed bioinformatic analysis. B.I., J.K., C.D.O. collected epidemiological and clinical data. A.L.W., C.B.F.V., I.M.O., R.E., S.L., P.L., A.V., A.P., M.T. performed the virus RNA concentration assays. N.D.G. supervised virus RNA extraction and concentration assays. A.C-M., M.C.M and A.J.M. processed and stored patient specimens, J.B.F., C.D.C., M.C. and S.F. assisted in patient recruitment. W.L.S. supervised and C.O. helped clinical data management. C.L. and A.I. drafted the manuscript. M.S., M.E., S.O. designed and conducted statistical analyses of the data. R.M., A.S., R.H., A.I. K. helped with interpretations of the data. All authors helped editing the manuscript. A.I. secured funds and supervised the project.

## Extended Data Figure/Table Legends

**Extended Table1: Basic Demographics for COVID-19 Cohort**. Unless otherwise noted, listed relative risks for mortality were not statistically significant. Moderate (Clinical Score1□3) and severe (Clinical Score 4□5) disease status were assigned as described in Methods. Percentages of sub□group (moderate or severe) are shown for each category with respective counts in parenthesis. Average age was calculated with accompanying sample standard deviation. Ethnicity and BMI were extracted from most recent electronic medical record (EMR) data. Select COVID[19 Risk Factors were scored by a clinical infectious disease physician. Presenting symptoms were recorded primarily through direct interview with patient or surrogate.

**Extended Data Fig**.**1: Age and BMI cohort distributions and Select Medications distributions:** Aggregated ages **(a)** and BMIs **(b)** were collected for moderate, severe, and fatal patients with COVID 19 and relative frequency histograms generated for comparison across disease sub-groups. Gaussian and lognormal distributions were fit through least squares regression and compared for goodness of fit through differential Akaike information criterion (AICc) comparison. All distributions were best described by a Gaussian model except for age in the “Severe” disease category, which was best modeled by a lognormal distribution. **(c)** Proportion of YNHH patients receiving hydroxycholorquine (HCQ), tocilizumab (Toci), methylprednisolone (Solu-medrol), and remdesivir (Rem) are shown, stratified by disease severity. **(d)** Medication and age adjustments for IL-6 and T cells count.

**Extended Data Fig. 2: Overview of cellular immune changes in COVID-19 patients**. Immune cell subsets were plotted **(a)** as a concentration of millions of cells per mL of blood or **(b)** as a percentage of a parent population. Each dot represents a separate time point per subject (HCW, n= 50; Moderate, n= 118; Severe, n= 41). Significance of comparisons were determined by Wilcoxon Rank-Sum Test and indicated as such: * p < 0.05, ** p< 0.01, *** p < 0.001, and **** p <0.0001.

**Extended Data Fig. 3: Overview cytokine and chemokines profiles of COVID-19 patients. (a)** Quantification of cytokines in the periphery plotted as Log10-transformed concentrations in pg/mL. Each dot represents a separate time point per subject (HCW, n= 47; Moderate, n= 124; Severe, n= 45). Significance of comparisons were determined by Wilcoxon Rank-Sum Test and indicated as such: * p < 0.05, ** p< 0.01, *** p < 0.001, and **** p <0.0001.

**Extended Data Fig. 4: Longitudinal cytokines and chemokines of COVID-19 patients. (a)** Log10-transformed cytokine concentrations plotted continuously over time according to the days of symptom onset for patients with moderate disease (n= 112) or severe disease (n= 39). The dotted green line represents the mean measurement from uninfected health care workers. Regression lines are indicated by the dark blue (moderate) or red (severe) solid lines. Associated linear regression equations, Pearson’s Correlation Coefficients, and significance are in pink (moderate) or dark blue (severe). 95% confidence intervals for the regression lines are denoted by the pink (moderate) or dark blue (severe) filled areas.

**Extended Data Fig. 5: T cell immune profiles in moderate and severe patients. (a)** CD4^+^ and **(b)** CD8^+^ T cell populations of interest, plotted as a percentage of parent populations, over time according to the days following symptom onset for patients with moderate disease (n= 118) or severe disease (n= 41). Each dot represents a distinct patient and time point arranged by intervals of five days until 25 days. The dotted green line represents the mean measurement from uninfected health care workers.

**Extended Data Fig. 6: (a)** Log10-transformed cytokine concentrations plotted continuously NP viral load (expressed as log10 genomic equivalents (GE)/ml) per within an individual patient and time point. Regression lines are indicated by the dark blue (moderate) or red (severe) solid lines for patients with moderate disease (n= 112) or severe disease (n= 39), respectively. Associated linear regression equations, Pearson’s Correlation Coefficients, and significance are in pink (moderate) or dark blue (severe). 95% confidence intervals for the regression lines are denoted by the pink (moderate) or dark blue (severe) filled areas. **(b)** Correlation map of highly correlated cytokines with NP viral load in patients with moderate (blue) or severe disease (red). Pearson’s Correlation Coefficients are indicated in grey, connecting the central node, NP viral load, with peripheral nodes; p-values for each correlation are indicated above each peripheral node. **(c)** Length of hospital stay plotted per patient against an individual’s baseline plasma cytokine measurements (<11 days from symptom onset), which were grouped according to high or low expression (>0.5-log difference). **(d)** Baseline plasma cytokine measurements for each patient who was either discharged from the hospital or expired during treatment for COVID-19. Significance of comparisons were determined by Wilcoxon Rank-Sum Test and indicated as such: * p < 0.05, ** p< 0.01, *** p < 0.001, and **** p <0.0001.

**Extended Data Fig. 7: Distribution of days from symptom onset stratified by collection time point**. Violin plots comparing the distributions of days from symptom for each patient ordered by sequential IMPACT study time points (1-8). Study time points 7 and 8 are represented by discrete points for the single patient collected at each. Violin plots display median values (solid line) and associated quartiles (dashed lines).

**Extended Data Fig. 8: Risk of ICU admission and death according to biomarkers levels**. Forest plots comparing the risk of death among ill patients. Each effect estimate represents an individual regression estimate with a Poisson family, log link, and robust variance estimation; each model accounts for repeated measures within one individual through the use of generalized estimating equations (GEE). Measurements are divided into three time-periods: 0-12 days after symptom onset, 12-19 days after symptom onset, and ≥20 days after symptom onset. If an individual had more than one measurement of a biomarker during any particular time period, we used the average of all values. Each model controls for participant age and sex.

**Extended Data Fig. 9: Gating strategies for immune cell Gating strategies for the key cell populations described in Fig. 1 b-c, Fig. 2 d-f, and in extended figures. (a)** Leukocyte gating strategy to identify lymphocytes, granulocytes, monocytes, pDCs, and cDCs. **(b)** T cell surface staining gating strategy to identify CD4 and CD8 T cells, TCR-activated T cells, Terminally-differentiated T cells, and additional subsets. **(c)** Intracellular T cell gating strategy to identify CD4 and/or CD8 T cells secreting TNF-α, IFN-γ, IL-6, IL-2, GranzymeB, IL-4, and/or IL-17.

